# Increasing HIV Testing Among Sexual and Gender Expansive Men in Kazakhstan: A Stepped-Wedge Randomized Trial of a Community-Level Intervention

**DOI:** 10.1101/2024.08.01.24311235

**Authors:** Elwin Wu, Yong Gun Lee, Vitaliy Vinogradov, Gulnara Zhakupova, Gaukhar Mergenova, Alissa Davis, Emily A. Paine, Timothy Hunt, Kelsey Reeder, Sholpan Primbetova, Assel Terlikbayeva, Caitlin Laughney, Mingway Chang, Baurzhan Baiserkin, Asylkhan Abishev, Marat Tukeyev, Sabit Abdraimov, Alfiya Denebayeva, Sairankul Kasymbekova, Galiya Tazhibayeva, Mashirov Kozhakhmet

**Author notes:** Correspondence concerning this article should be addressed to Elwin Wu, Social Intervention Group, Columbia University School of Social Work, 1255 Amsterdam Avenue, New York, NY 10027. USA.

## Abstract

**Importance:** HIV transmission in Kazakhstan has increased among men who have sex with men (MSM) and transgender and nonbinary people who have sex with men (TSM), driven by low HIV testing rates.

**Objective:** To determine if the *PRIDE in HIV Care* intervention had a community effect of increasing HIV testing among MSM and TSM in Kazakhstan.

**Design:** We employed a stepped-wedge, cluster-randomized controlled trial with MSM and TSM community members recruited from three cities in Kazakhstan: Almaty, Astana, and Shymkent. We collected serial cross-sectional data where community members completed one assessment between 21 August 2018, and 30 March 2022.

**Setting:** We collected data from 629 MSM and TSM among the study cities. Community respondents were recruited from real-world (e.g., NGOs, bars, clubs) or virtual sites (e.g., social media, apps) where MSM and TSM in each of the three cities were known to frequent.

**Participants:** Eligibility criteria for community respondents were: (1) ≥18 years old; (2) identifying as male at any point in life or being assigned male at birth; (3) having consensual sex with another man in the past 12 months; (4) engaging in binge drinking (i.e., ≥5 drinks in a 2 hour period), illicit use of drugs, or both in the past 90 days; and (5) residing in one of the three study cities.

**Intervention:** The *PRIDE in HIV Care* intervention is a theory-driven “crowdsourcing and peer-actuated network intervention” designed to amplify community members’ successes and resilience via “influencers” who can strengthen and impart benefit to their networks and community.

**Main outcome measures:** Received an HIV test in the prior six months.

**Results:** There was a statistically significant increase in odds of recent HIV testing for every additional month the intervention was implemented in a respondent’s city (*AOR*=1.08, *95% CI*=1.05-1.12; *p*<.001).

**Conclusions:** The *PRIDE in HIV Care* intervention appears to be efficacious in enacting a community wide increase—i.e., promoted HIV testing among those who did not go through the intervention itself—in HIV testing among MSM and TSM.

**Trial Registration:** This trial is registered with clinicaltrials.gov (NCT02786615).

**Funding:** National Institute on Drug Abuse (NIDA), grant number R01DA040513.

**KEY POINTS:** *Question:* Does the *PRIDE in HIV Care* intervention exert a community effect of increasing HIV testing among men who have sex with men (MSM) and transgender and nonbinary people who have sex with men (TSM) in Kazakhstan?

*Findings:* We employed a stepped-wedge, cluster-randomized controlled trial among three cities in Kazakhstan. There was a statistically significant increase in odds of recent HIV testing for every additional month the intervention was implemented in a respondent’s city.

*Meaning:* The intervention increased HIV testing among MSM and TSM in Kazakhstan who had not directly received the intervention, providing support for a community-wide impact.

## INTRODUCTION

Public health efforts to end the HIV epidemic have successfully reduced HIV incidence and improved HIV care outcomes across many national contexts. In Kazakhstan, however, HIV incidence has increased 88% from 2010 to 2021—the 7th highest increase in the world—and the number of people living with HIV more than doubled.^1,2^ This trend is accelerated among gay, bisexual, and other men who have sex with men (MSM), who experienced an increase in prevalence from 1·2% in 2013^3^ to 6·5% in 2020.^1^ Additional studies suggest higher prevalence among some sub-groups of MSM and transgender and nonbinary people who have sex with men (TSM),^4–6^ such as those who use substances and/or engage in binge drinking (15.6%).^6^ In 2019, it was estimated that only 30% of MSM living with HIV knew their status,^7^ signaling a great need to increase HIV testing.

MSM and TSM in Kazakhstan face significant barriers to HIV testing and engagement in the HIV care continuum. Evidence suggests that HIV stigma as well as stigma arising from homophobia and transphobia are pervasive and impede access to care.^6,8–13^ Resultant internalized homophobia and transphobia compromise psychosocial wellbeing and have been associated with lower rates of HIV testing among MSM and TSM in Kazakhstan.^6,8,14^

Despite the demonstrated need, evidence-based HIV preventive interventions involving MSM and TSM in Kazakhstan are scarce. To address this gap, we developed and tested an intervention for increasing the number of MSM and TSM engaged in the HIV care continuum in Kazakhstan. We built upon more than two decades of evidence-based social network interventions,^15^ demonstrating their flexibility and utility in disseminating HIV information among peers and leveraging social support to improve HIV outcomes. Given the longstanding oppression and marginalization of MSM and TSM in Kazakhstan, we buttressed the peer influence and social network approach with community empowerment.^16^ In particular, viewing MSM and TSM as experts and catalysts of change, we utilized crowdsourcing, the process of engaging the public to develop and share solutions.^17,18^ Finally, we drew upon social marketing principles and practices^19–27^ to optimize promotion reach and impact within networks.

Synthesizing across these approaches, we conceptualized and developed the *Peer Reach and Influencer-Driven Engagement in the HIV Care Continuum (PRIDE in HIV Care)* intervention as a crowdsourcing and peer-actuated network intervention. We then employed a stepped-wedge randomized clinical trial that utilized a serial cross-sectional data collection and analysis to test its efficacy on increasing broader community-wide HIV testing. Specifically, we hypothesized that the odds of MSM and TSM community members (who were not directly participating in the intervention) having a recent HIV test would increase after the implementation of the intervention in their city of residence.

## METHODS

### Trial Design

This study was an open-label stepped-wedge randomized trial focused on a target population of MSM and TSM in Kazakhstan engaged in substance use; the trial period covered 21 August 2018 to 30 March 2022. Because the social networks of individuals are not known in advance—hence cannot be reliably assigned to a single experimental condition—and the peer influence mechanism can exert an influence over some geographical distance, we utilized a design in which experimental control and random assignment were performed at the city level in three geographically disparate cities: Almaty, Astana, and Shymkent. These cities were chosen based on (1) being among the cities with the highest prevalence and incidence of HIV in the country, and (2) having physical and digital infrastructures to access MSM and TSM residing there.^28^ Figure 1 presents a CONSORT diagram depicting the major aspects of trial design and performance. The trial began with a 6-month “pre-implementation” period in which all cities had no intervention delivered/available. Study steps were planned to be six months in duration, with intervention delivery beginning in one new city each subsequent step until the intervention was delivered/available in all three cities (total of 18 months). Data collection and analysis utilized a serial cross-sectional design; all respondents were unique individuals.

**Figure 1:**
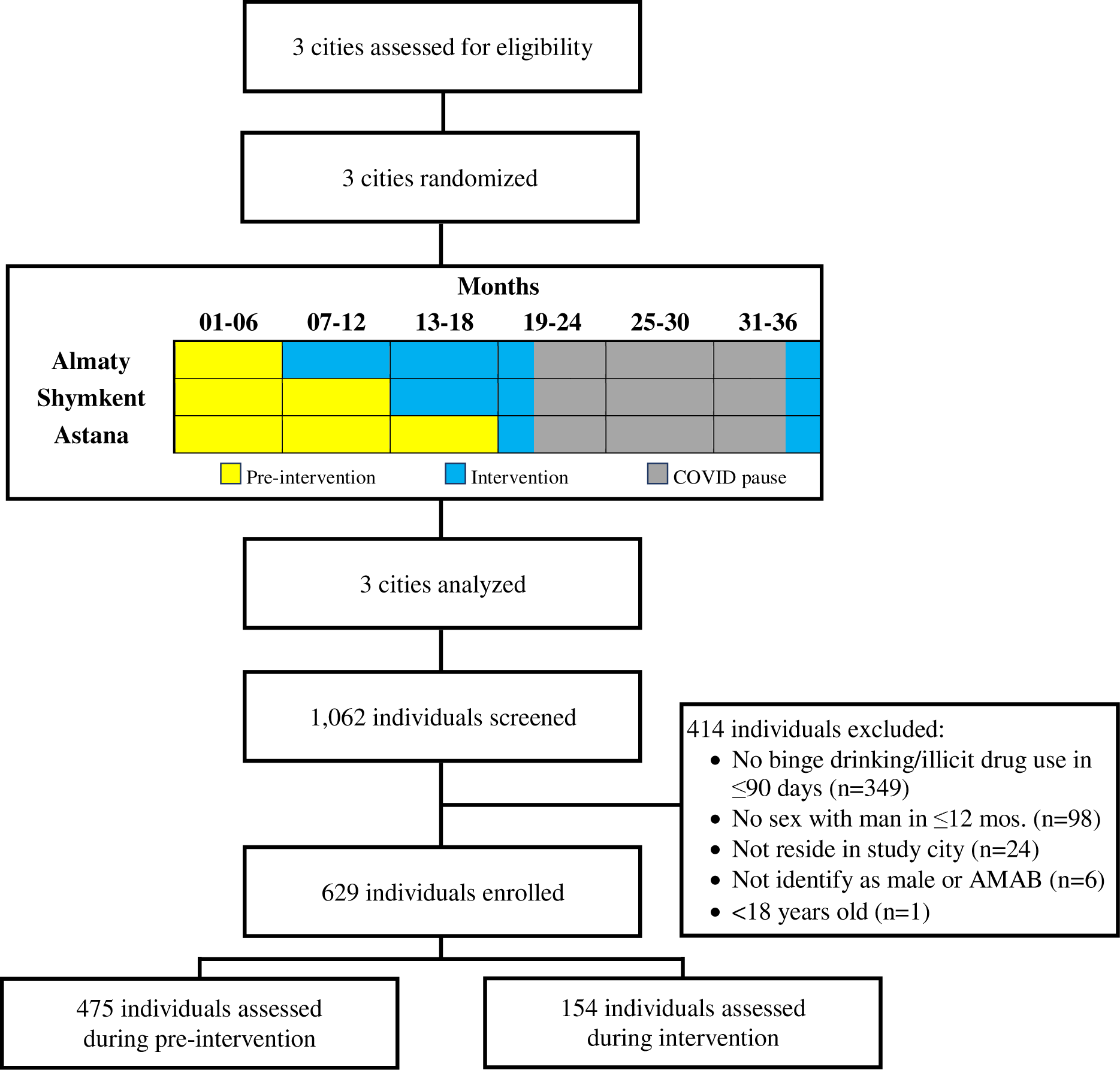
CONSORT Diagram.

Two unexpected changes to the trial design occurred due to the COVID-19 pandemic. First, COVID-19 emergence and transmission mitigation measures resulted in a loss of recruitment time and, consequently, a lower-than-targeted sample size. Second, intervention delivery shifted from an in-person, group-based modality to a remotely delivered, individual modality. The pause began in March 2020, initially involving a halt in participant-facing study activities, while intervention delivery protocols were revised to be conducted remotely (e.g., via internet or digital telecommunications apps); the study’s Institutional Review Boards (IRBs) approved remote intervention delivery in December 2020. In-person activities, specifically the main assessment—hence enrollment—were allowed to recommence in November 2021.

### Intervention

The *PRIDE in HIV Care* intervention was a novel crowdsourcing and peer-actuated network intervention designed to promote engagement in the HIV care continuum among MSM and TSM in the community. The intervention drew upon Social Cognitive Theory (SCT)^29^, social marketing principles,^19^ and adult learning theories^30^ to focus on facilitating intervention recipients become “influencers” for HIV testing and treatment among MSM and TSM community members.

Figure 2 presents the formal logic model for the intervention. Activities were specifically designed to target intervention mediators and key clinical processes (e.g., attendance motivation, safety). For instance, crowdsourcing prompted intervention recipients to share strategies for effectively engaging in the HIV care continuum, including identifying MSM/TSM-friendly providers, coping with stigma, and fostering or maintaining motivation for testing. Intervention activities also encouraged recipients to use contemporary digital marketing approaches (e.g., influencer marketing, viral marketing) in social marketing. To aid in the application of social marketing concepts, the intervention had recipients consider the following (with example prompts):

- Behavior (What is the desired behavior? e.g., getting an HIV test)
- Location (Where might the behavior be performed?)
- Audience (Who are you targeting?)
- Strategy (How will you promote the behavior? e.g., tone of messaging)
- Tools (What do you need to execute your marketing strategy? e.g., social media account).

**Figure 2:**
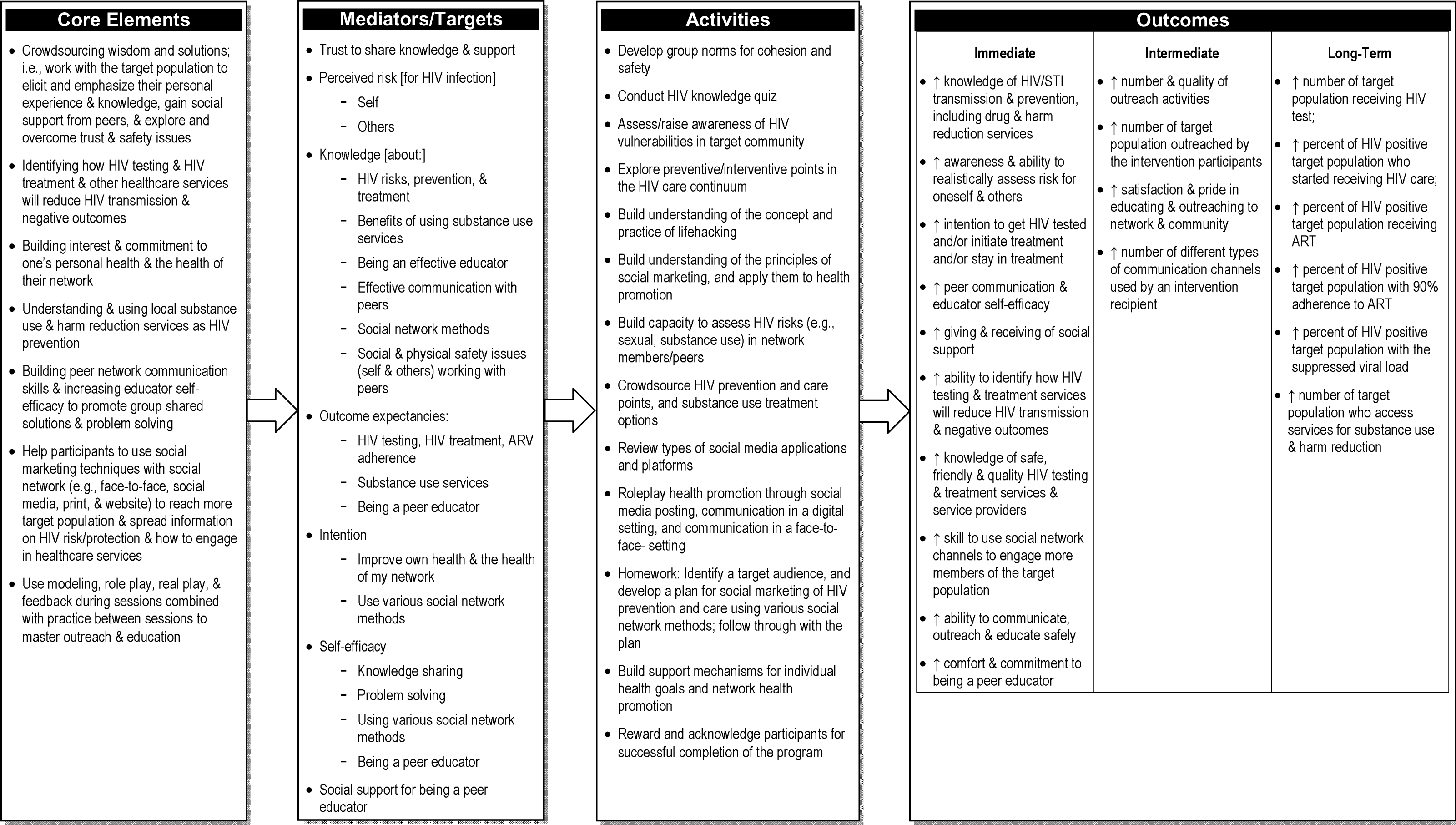
Logic model for the *PRIDE in HIV Care* intervention.

Prior to the COVID-19 study interruption, the intervention was delivered as a single one-on-one orientation session followed by four facilitator-led group sessions. During the COVID-19 study pause, the intervention was adapted to be delivered remotely in a one-on-one fashion. This allowed the study to resume in accordance with COVID-19 mitigation measures/requirements. The adaptation method to ensure rigor and reproducibility have been published previously.^31^

Figure 3 presents representative examples of materials developed (i.e., crowdsourced) and used by intervention recipients during the trial to influence and assist MSM and TSM community members in their city to engage in HIV testing, prevention, and care.

**Figure 3:**
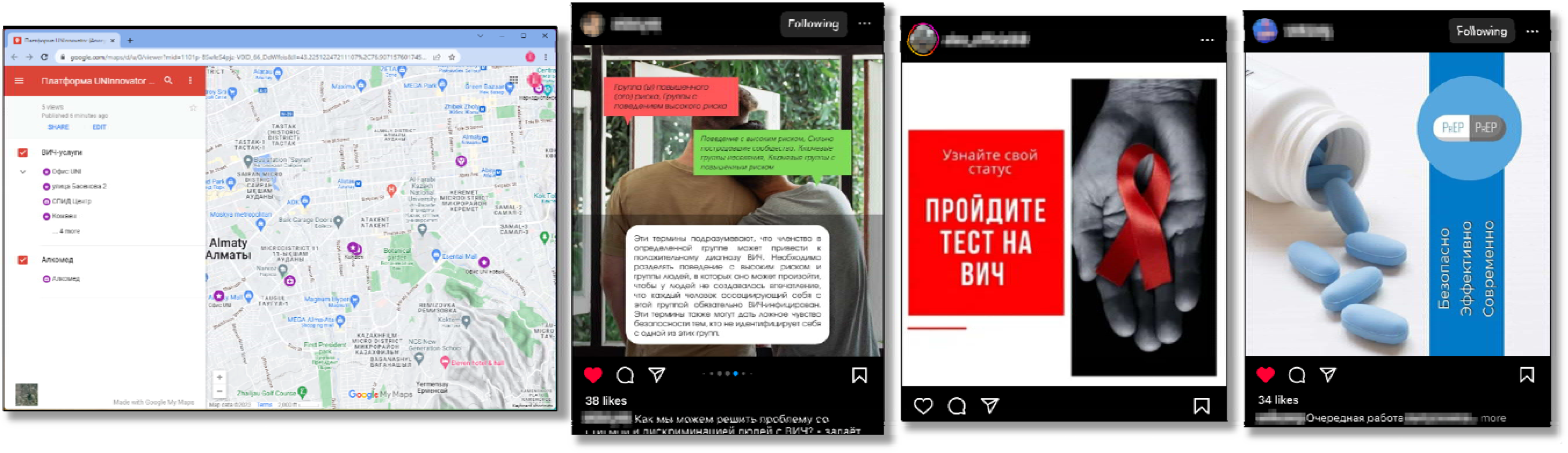
Sample individual and crowdsourced items developed and social marketed by *PRIDE in HIV CARE* intervention recipients. From left to right: a crowdsourced map sharable for users to add and “pin” local resources for MSM and TSM; social media post promoting well-being of MSM (Red box: *“High risk group(s), groups with high-risk behavior”*, Green box:*“High risk behavior, community that suffered greatly, key population groups, key groups at increased risk”*, White box: *“These terms imply that membership in a particular group can lead to a positive HIV diagnosis. It is important to distinguish between high-risk behavior and the groups of people in whom it may occur, so that people do not get the impression that every person who associates with this group must be infected with HIV. These terms can also provide a false sense of security to those who do not identify with one of these groups.”)*; social media post promoting HIV testing (*“Know your status, get tested for HIV”)*; and a social media post promoting PrEP (*“PrEP – Safe, Effective, Modern”*); (Note: images have been edited, solely to preserve anonymity)

### Community Respondents

We underscore that the intervention was primarily designed to exert an impact in the community rather than spur individual change among intervention recipients. Thus, we assess primary outcomes among community respondents who were not involved in the intervention at the time of assessment.

Starting six months before the trial, research staff recruited community respondents at real-world (e.g., NGOs, bars, clubs) and virtual (e.g., Instagram, Grindr) sites where MSM and TSM were known to frequent.^28^ Respondents also referred people from their social networks to the study team as potential respondents.

Eligibility criteria for community respondents were based on self-report and included: (1) ≥18 years old; (2) identifying as male at any point in life or assigned male at birth (AMAB); (3) having consensual sex with another man in the past 12 months; (4) engaging in binge drinking (i.e., ≥5 drinks in a 2 hour period), illicit use of drugs, or both in the past 90 days; and (5) residing in one of the three study cities. Individuals were excluded if language and/or cognitive abilities prohibited providing informed consent. Eligibility was determined via a brief computer-assisted structured interview (CASI) administered by a trained interviewer.

### Randomization

Six months prior to the start of the trial, the investigative team used a computerized random number generator to determine the order in which cities would enter the implementation phase in the stepped-wedge design. Respondents were not informed about the implementation status of their city at the time of data collection.

### Data Collection and Measurement

Data collection followed a serial cross-sectional design. Each respondent completed a survey at a single timepoint administered in Kazakh or Russian (at the preference of the respondent) by trained interviewers. The survey consisted of questionnaires used in prior studies focused on HIV and/or substance use by the investigative team, including studies with MSM.^32^

#### Primary Outcome

Respondents reported if they ever had an HIV test (prior to this study) and the time (month and year) of their most recent HIV test. For the primary outcome of this study, respondents who had received an HIV test in the past six months were coded 1=yes, 0=no.

#### Covariates

Sociodemographic data included age, sex assigned at birth, current gender identity, sexual orientation, marital status, employment, and monthly income.

Respondents self-reported lifetime binge drinking and use of marijuana/cannabinoids, heroin/opioids, stimulants, cocaine, hallucinogens or psychedelics, inhalants, and club drugs. If a respondent indicated lifetime use, they were asked about recent use (i.e., in the past 90 days).

Biological assays for HIV and STIs were conducted immediately after the survey. Community respondents self-collected urine and rectal swab specimens, which were shipped to certified laboratories in each city for testing of *Chlamydia trachomatis* and *Neisseria gonorrhea* (AmpliSense molecular/DNA amplification assay). Finger prick blood samples were used for syphilis testing (Alere Determine Syphilis TP rapid screening test). For respondents with reactive results, venous blood was collected for confirmatory testing using a non-treponemal test (test of Venereal Diseases Research Laboratory [VDRL]) and a second treponemal test (*Treponema pallidum* particles agglutination [TPHA]) when needed. The study also provided HIV testing, but the study-provided HIV test was excluded from the analysis approach for the primary outcome of recent HIV testing.

### Statistical Methods

#### Primary Outcome

The primary outcome for the study was recent HIV testing (i.e., receiving an HIV test in ≤ six months). Respondents were instructed to not consider the study-provided HIV test. As noted earlier, the intervention was designed to make an impact on engagement in the HIV care continuum *among MSM and TSM in the community* (as opposed to increasing such behaviors among intervention recipients themselves). Thus, efficacy analyses focused on HIV testing behavior prior to a respondent being able receive the intervention to become an HIV care continuum influencer.

#### Sample Size

Power analyses based on the original study design predicted that 1,000 participants would provide 80% statistical power for the step in the HIV care continuum having the hardest-to-detect outcome, which was ART adherence among those living with HIV. The unavoidable pause in study activities due to COVID-19 resulted in a loss of about a third of the planned recruitment time. We enrolled 629 participants, which was proportionate of the actual vs. planned recruitment time; this was still adequately powered for HIV testing but not for the other points in the HIV care continuum.

#### Inferential hypothesis testing

For the main analysis, we used a Generalized Linear Model with a logit link function to test the hypothesis that the primary outcome of recent HIV testing would increase as a function of time (measured in months) since the intervention began to be implemented in a respondent’s city. Models accounted for clustering within cities using random slopes and intercepts. To account for secular trends, the initial model included time-in-months from start of study as a constant, and time-in-months since the intervention was implemented in the respondent’s city at the time of data collection from the respondent (for periods before intervention implementation in a city, the value of this variable was set to zero). This “bent stick” model has been used in prior efficacy tests of stepped-wedge randomized trials.^33^ The final model added covariance adjustment for self-reported sociodemographics, recent binge drinking and illicit use of drugs, and STI status (independently for Chlamydia, Gonorrhea, Syphilis) determined via biological assay.

Post hoc analyses were conducted using the same analytic approach described above to (1) restrict data to the period before COVID-19 and, hence, before the change of the intervention delivery to the remote modality; and (2) gain preliminary insight into the impact of the substance use criterion on the generalizability of the sample/findings.

### Research Ethics and Review

All respondents provided informed consent at the start of screening and assessment visits. Respondents were compensated with gift cards with 2000 and 6000 Kazakhstan Tenge (∼$5 and ∼$15USD) for completion of the screening and main assessment respectively. All study procedures were approved by the Institutional Review Boards at Columbia University and Kazakhstan National University.

## RESULTS

We conducted 1062 screening interviews across the three cities (*n*=437, 330, and 295 in Almaty, Astana, and Shymkent respectively). Overall, 648 individuals screened eligible for the study (Figure 1). We enrolled 629 (97%) of study-eligible individuals, which constitutes the analytic sample for the primary outcome hypothesis testing.

There were significant differences for most of the sociodemographic characteristics by city (Table 1). Shymkent had the highest proportion of participants who preferred to communicate in Kazakh, were transgender, bisexual/pansexual/etc., married, did not complete a high school education, and employed part-time or unemployed.

**Table 1:** Sociodemographic and clinical characteristics (N=629)

|  | **Total sample**<br>(N=629) | **Almaty**<br>(n=245) | **Astana**<br>(n=194) | **Shymkent**<br>(n=190) | **p-value**<br>for difference<br>among cities |
| --- | --- | --- | --- | --- | --- |
| **Age (yrs.)** $\bar{x}$ (SD) | 29.0 (9.0) | 29.8 (7.9) | 27.1 (7.0) | 29.9 (8.6) | <.001 |
| **Preferred language** *n* (%) |  |  |  |  |  |
| Russian | 553 (88%) | 238 (97%) | 182 (94%) | 133 (70%) | <.001 |
| Kazakh | 76 (12%) | 7 (2.9%) | 12 (6.2%) | 57 (30%) |  |
| **Cisgender man** *n* (%) | 560 (89%) | 217 (89%) | 179 (92%) | 164 (86%) | .17 |
| **Sexual orientation** *n* (%) |  |  |  |  |  |
| Gay/homosexual/etc. | 323 (51%) | 134 (55%) | 116 (60%) | 73 (38%) | .002 |
| Bisexual/pansexual/etc. | 268 (43%) | 101 (41%) | 66 (34%) | 101 (53%) |  |
| Straight/heterosexual | 11 (1.7%) | 3 (1.2%) | 3 (1.5%) | 5 (2.6%) |  |
| Other | 27 (4.3%) | 7 (2.9%) | 9 (4.6%) | 11 (5.8%) |  |
| **Marital status** *n* (%) |  |  |  |  |  |
| Single, never married | 496 (79%) | 202 (82%) | 152 (78%) | 142 (75%) | <.001 |
| Married | 49 (7.8%) | 12 (4.9%) | 9 (4.6%) | 28 (15%) |  |
| No longer with spouse | 55 (8.7%) | 26 (11%) | 11 (5.7%) | 18 (9.5%) |  |
| Other | 29 (4.6%) | 5 (2.0%) | 22 (11%) | 2 (1.1%) |  |
| **Education**<sup>a</sup> *n* (%) |  |  |  |  |  |
| Less than high school | 41 (6.7%) | 6 (2.5%) | 11 (6.1%) | 24 (12.6%) | <.001 |
| High school to some college | 281 (46%) | 106 (43%) | 79 (44%) | 96 (51%) |  |
| Baccalaureate or higher degree | 292 (48%) | 132 (54%) | 90 (50%) | 70 (35%) |  |
| **Employment** *n* (%) |  |  |  |  |  |
| Working full-time | 331 (53%) | 134 (55%) | 106 (55%) | 91 (48%) | .001 |
| Working part-time | 153 (24%) | 65 (27%) | 33 (17%) | 55 (29%) |  |
| Student | 67 (11%) | 16 (6.6%) | 34 (18%) | 17 (8.9%) |  |
| Unemployed | 76 (12%) | 29 (12%) | 20 (10%) | 27 (14%) |  |
| **Monthly income (KZT × 1000)** $\bar{x}$ (SD) | 192 (332) | 189 (192) | 194 (179) | 192 (534) | .99 |
| **Substance use (past 90 days)** *n* (%) |  |  |  |  |  |
| Binge drinking | 532 (85%) | 205 (84%) | 160 (83%) | 167 (88%) | .30 |
| Illicit use of drugs | 260 (41%) | 111 (45%) | 77 (40%) | 72 (38%) | .25 |
| **Sexually transmitted infection** *n* (%) |  |  |  |  |  |
| Chlamydia | 111 (18%) | 34 (17%) | 38 (20%) | 39 (22%) | .39 |
| Gonorrhea | 48 (7.6%) | 20 (9.9%) | 10 (5.2%) | 18 (10%) | .15 |
| Syphilis | 128 (20%) | 42 (17%) | 46 (24%) | 40 (21%) | .22 |

<sup>a</sup> N= 614 (15 missing observations due to “refuse to answer”)

Among this sample, 254 (40%) reported having received an HIV test in the past six months. These testing rates differed significantly (*p*<·001) by city, with 112 (46%), 91 (47%), and 51 (27%) of the participants from Almaty, Astana, and Shymkent respectively who underwent HIV testing in the prior six months.

### Efficacy Outcomes

Results from the primary outcome analyses (Table 2) indicate a statistically significant increase in odds of recent HIV testing for every additional month the intervention was implemented in a respondent’s city (*AOR*=1.08, *95% CI*=1.05-1.12; *p*<.001) and this offsets the statistically significant estimated negative trend over time in HIV testing (*AOR*=0.95, *95% CI*=0.93-0.97; *p*<.001). These relationships remain significant and within their *95% CI*s with covariance adjustment for sociodemographic factors, substance use behaviors, and STI status.

**Table 2:** Adjusted Odds Ratios of Recent HIV Testing (primary outcome)

|  | *AOR (95% CI)* | *p-value* |  | *AOR (95% CI)* | *p-value* |
| --- | --- | --- | --- | --- | --- |
| **Mos. of intervention implementation** | 1.08 (1.05-1.12) | <.001 |  | 1.07 (1.04-1.11) | <.001 |
| **Calendar time (mos.)** | 0.95 (0.93-0.97) | <.001 |  | 0.96 (0.95-0.98) | <.001 |
| **Constant** | 0.93 (0.67-1.31) | .67 |  | 0.69 (0.07-7.41) | .76 |
| **Age (yrs.)** |  |  |  | 0.99 (-.95-1.03) | .58 |
| **Preferred language** |  |  |  |  |  |
| Russian |  |  |  | 2.17 (1.47-3.20) | <.001 |
| Kazakh |  |  |  | ref. |  |
| **Cisgender man** |  |  |  | 1.62 (1.04-2.53) | .03 |
| **Sexual orientation** |  |  |  |  |  |
| Gay/homosexual/etc. |  |  |  | ref. |  |
| Bisexual/pansexual/etc. |  |  |  | 0.43 (0.29-0.64) | <.001 |
| Straight/heterosexual |  |  |  | 0.51 (0.39-0.68) | <.001 |
| Other |  |  |  | 0.87 (0.28-2.69) | .99 |
| **Marital status** |  |  |  |  |  |
| Single, never married |  |  |  | ref. |  |
| Married |  |  |  | 0.99 (0.55-1.77) | .96 |
| No longer with spouse |  |  |  | 0.72 (0.21-2.50) | .61 |
| Other |  |  |  | 0.93 (0.25-3.48) | .91 |
| **Education** |  |  |  |  |  |
| Less than high school |  |  |  | 0.42 (0.23-0.77) | .01 |
| High school to some college |  |  |  | 1.02 (0.60-1.75) | .93 |
| Baccalaureate or higher degree |  |  |  | ref. |  |
| **Employment *n* (%)** |  |  |  |  |  |
| Working full-time |  |  |  | ref. |  |
| Working part-time |  |  |  | 1.07 (0.70-1.62) | .76 |
| Student |  |  |  | 1.23 (0.47-3.23) | .67 |
| Unemployed |  |  |  | 1.10 (0.40-3.02) | .86 |
| **ln (Monthly income)** |  |  |  | 1.02 (0.88-1.18) | .82 |
| **Substance use (past 90 days)** |  |  |  |  |  |
| Binge drinking |  |  |  | 0.51 (0.39-0.66) | <.001 |
| Illicit use of drugs |  |  |  | 1.24 (1.21-1.27) | <.001 |
| **Sexually transmitted infection** |  |  |  |  |  |
| Chlamydia |  |  |  | 0.73 (0.46-1.15) | .17 |
| Gonorrhea |  |  |  | 1.31 (0.64-2.71) | .46 |
| Syphilis |  |  |  | 1.64 (1.25-2.14) | <.001 |

Adjusted odds ratios (*AORs*) and associated 95% confidence intervals (*95% CIs*) for the primary outcome measure of receiving an HIV test in the past 6 months estimated using multilevel logistic regression models with random slopes and intercepts for each study city.

#### Ancillary analyses

We conducted an ancillary analysis restricting data to those collected pre-COVID-19; the statistical significance remained unchanged. As an exploratory analysis regarding the impact of the substance use eligibility criterion, we analyzed screening data for MSM and TSM regardless of the substance use criterion (i.e., including MSM or TSM who did not engage in binge drinking nor illicit drug use in the past 90 days) but met all the other eligibility criteria, albeit with covariance adjustment with the variables available at screening (age, income, sexual orientation, being cisgender, marital status, preferred language, employment, and recent binge drinking and illicit use of drugs); the odds of having a recent HIV test among this larger sample (*N*=849) of MSM and TSM for each month the intervention was implemented in the respondent’s city was statistically significant in a beneficial direction (*AOR*=1.08, *95% CI*=1.06-1.10; *p*<.001); this effect size offsets the statistically significant negative trend over time in HIV testing (*AOR*=0.96, *95% CI*=0.95-0.96; *p*<.001).

## DISCUSSION

Results support *PRIDE in HIV Care* as an efficacious behavioral intervention that can increase HIV testing among MSM and TSM communities in Kazakhstan. Of note and particular value, the intervention was designed and assessed to have a community-level effect: *PRIDE in HIV Care* can prompt behavior change among individuals who never directly received the intervention.

The intervention was designed such that intervention effects would diffuse out through a recipient’s social networks. The stepped-wedge randomized trial design accommodated for social networks within a city. It has limited ability to control for secular and external events that exert non-linear temporal trends. Contamination across cities is still possible, especially given digital social media which can have a wide geographical reach. Yet the use of three socioeconomically varied cities geographically dispersed across the country is a strength. The COVID-19-driven pause resulted in several unavoidable changes: a decrease in sample size and loss of statistical power to detect secondary outcomes (e.g., receiving ART, achieving viral suppression); and intervention delivery modality being confounded with time (i.e., all intervention delivery starting in January 2021 was remote). However, even with the smaller-than-planned sample size, we believe this is still considerably the largest sample of MSM and TSM in Kazakhstan reported in the behavioral science literature to date.

Most of these limitations would result in decreasing the detectable effect and, thus, increase Type II error. However, we were able to reject the null hypothesis for the primary outcome, indicating that the substantial strengths of our study and the stepped-wedge design outweigh the limitations. Given that HIV testing represents the greatest gap in the HIV care continuum for MSM and TSM in Kazakhstan,^28^ our findings have significant implications for future HIV programs and research.

### Conclusions

This clinical trial supports the addition of *PRIDE in HIV Care* to the set of evidence-based HIV preventive interventions and advances evidence-based community-level, peer HIV prevention in other ways. A new community-level intervention is a noteworthy advance given the difficulties and accompanying scarcity of rigorous trials designed to change the social milieu in ways that lead to HIV-protective behavior.^34,35^ This intervention also uses contemporary digital social marketing, virtual social networks, and social media, whose importance took on greater significance with disruptions to traditional intervention delivery venues due to COVID-19 mitigation protocols that disrupted face-to-face delivery. With respect to cultivating peers for promotion and/or social marketing, the crowdsourcing approach reduces the necessity of ethnographic and social network mapping steps needed to identify popular and socially influential members of the target population; these steps are not only time and resource intensive, but also may prove particularly challenging for key populations experiencing oppression. Crowdsourcing also ensures the ways to overcome challenges are ecologically valid for the local service system, sociocultural milieu, and safety considerations. These benefits are buttressed by *PRIDE in HIV Care*’s social marketing skill enhancement, which has been updated for current social trends (e.g., influencers) and technologies (e.g., digital social media). Remote delivery of the intervention also offered an important avenue for enhancing scale-up and increasing its dissemination and reach. Given that *PRIDE in HIV Care* strengthens and amplifies the local supports and strengths within a community, we hope that this intervention provides a valuable program and template for community empowerment in addressing future psychosocial and health issues.

## Contributors

EW, TH, SP, AT, and BB contributed to study conceptualization, funding acquisition, methodology, and investigation. YGL, VV, GZ, GM contributed to project administration and data curation. EW, YGL, VV, GZ, GM, EAP, TH, KR, SP, and AT provided supervision of staff and oversight of key areas of the study. EW and MC led the formal analyses. EW wrote the initial draft and all authors contributed to manuscript review and editing. All authors contributed to interpretation of the work and final approval of this manuscript.

## Data Availability

The data that support the findings of this study are available from the corresponding author, Dr. Elwin Wu, upon reasonable request.

## Acknowledgements

In addition to the co-authors, this work was only made possible by the research staff and allies who showed tremendous dedication, skill, and sensitivity—Gassan Akhmedov, Karina Alipova, Farruh Aripov, Olga Balabekova, Daniyar Bekishev, Dilara Belkesheva, Valeriya Davydova, Ferangiz Hasanova, Altynay Kambekova, Sultana Kali, Saltanat Kuskulova, Aitkul Nazarova, Syrym Omirbek, Anna Ryl, Svyatoslav Suslov, Aizhan Toleuova, Aidar Yelkeyev, and Saida Yessenova.—and, most importantly, the time and trust given by the participants and many courageous MSM and TSM in Kazakhstan. We also thank Dr. Jeremy Sugarman for his invaluable consultation throughout the study. The findings and conclusions in this report are those of the authors and do not necessarily represent the views of the NIDA or the Columbia University School of Social Work.

## Registration

This study is registered with clinicaltrials.gov, number NCT02786615.

## Protocol

Protocol materials are available upon request.

